# Increased risk of fungal infection detection in women using menstrual cups vs. tampons: a cross-sectional study

**DOI:** 10.1101/2021.12.10.21267584

**Authors:** Nicolas Tessandier, Ilkay Başak Uysal, Baptiste Elie, Christian Selinger, Claire Bernat, Vanina Boué, Sophie Grasset, Soraya Groc, Tsukushi Kamiya, Massilva Rahmoun, Bastien Reyné, Noemi Bender, Marine Bonneau, Christelle Graf, Vincent Tribout, Vincent Foulongne, Jacques Ravel, Tim Waterboer, Christophe Hirtz, Ignacio G Bravo, Jacques Reynes, Michel Segondy, Carmen Lia Murall, Nathalie Boulle, Samuel Alizon

## Abstract

**Objective:** To determine if the use of menstrual cups rather than tampons is associated with more or less health risk.

**Design:** Analysing biological, demographic, and behavioural data in a cohort of women who reported using mostly tampons (*n* = 81) or menstrual cups (*n* = 22).

**Setting:** A cross-sectional analysis using the inclusion data of a single centre longitudinal study.

**Population:** 149 women from 18 to 25 years old living in the area of Montpellier (France) who reported having at least one new sexual partner over the last year.

**Methods:** Statistical modelling (mainly binomial regression models and factor analyses of mixed data).

**Main Outcome Measures:** Self-reported data from questionnaires (fungal infection, urinary tract infection, stress level) and biological data (HPV screening, vaginal microbiota profiling, circulating antibodies titration, and local cytokine concentrations).

**Results:** We identify an increased risk of reporting fungal infections for women using menstrual cups over tampons. We do not detect significant differences in terms of vaginal microbiota composition or local cytokines expression profile but find that women fall into two different clusters in a factor analysis of mixed data depending on the type of menstrual product they use more (cups or tampons).

**Conclusions:** These results point to potential health risks in the use of menstrual cups and differences in local vaginal environments. In-depth studies are needed to better understand potential associations between menstrual product use and women’s health.

**Funding:** European Research Council (EVOLPROOF, grant 648963)

**Ethics:** The PAPCLEAR study ClinicalTrials.gov identifier is NCT02946346.

**Tweetable abstract:** A cross-sectional study finds a significant association between menstrual cup use and fungal infection risk.

## Introduction

Menstruation is associated with reproductive health, but it also has implications beyond physical health, as recognised by the emergent concept of menstrual health [1]. Further-more, the risks associated with menstrual poverty [2] are increasingly recognised and include for instance bacterial vaginosis [3] or reproductive tract infections [4].

The vaginal microbiota is a crucial component of women’s health throughout their lives [5]. It could play a key role in preventing diseases such as yeast infections, urinary tract infections (UTI), and STIs, especially HIV [6]. Contrarily to the gut microbiota, the vaginal microbiota is associated with relatively few, clearly-defined, community state types (CST), most of which are dominated by one or two species of *Lactobacillus* bacteria [7]. The CST IV stands out because of its high diversity and prevalence of anaerobic bacteria with, therefore, an association with dysbioses. For these reasons, it could be associated with menstrual health.

The immune system (both innate and adaptive) elements of the vaginal environment have evolved to meet the special challenges that are associated with the female reproductive tract [8]. This equilibrium is tightly maintained by both sex hormones throughout the menstrual cycle and microbiota to respond to the challenges of bacteria, yeast, and viruses without interfering with events that surround conception[9]. Perturbations in either may impede the efficiency of the immune system to control pathogens and significantly compromise women’s health.

Among the variety of products used during the menses, menstrual cups are perceived as a safe, practical, economical, and ecologically-friendly alternative to tampons and sanitary pads [10]. The majority of women using them report wanting to continue to use them both in high and low-income settings, showing a good level of acceptability [11, 10]. Nevertheless, there have been some reported cases of toxic shock syndrome, renal colic, and allergies associated with menstrual cup usage [10]. Moreover, higher levels of *Staphylococcus aureus* growth have been reported in menstrual cups compared to tampons [12]. However, the number of case studies is limited and we know little about potential health risks associated with cup usage compared to other menstrual hygiene products [10].

Our objective is to test whether the type of menstrual hygiene products can shape the vaginal immunological and microbiological environment in a way that could affect women’s health. To this end, analysed biological, demographic, and behavioural longitudinal data from 149 women, in the context of the PAPCLEAR clinical study on human papillomavirus (HPV) infections [13]. We focused in particular on the local impact of the different product use and, using statistical modelling, identified profile differences depending on the type of menstrual product used.

## Materials and methods

### Cohort description and data curation

The participants of this analysis were 149 young women from the PAPCLEAR longitudinal clinical study which started in 2016 and was finalised in 2020. The women were aged between 18-25 years, mainly students, from the area of Montpellier (France) and their human papillomavirus status, other genital infections, immunological responses (antibodies and cytokines), behaviours were followed up for two years [13].

We selected participants who reported using tampons or cups for menstrual products, and for whom detailed cytokine profiles (see below), microbiota metabarcoding data, and antibody data at the inclusion visit were available. This amounts to *n* = 103 women.

We assigned tampon or menstrual cup categories when a participant reported using either type of menstrual product over 75% of the time over the whole duration of the study. There was no difference in follow-up duration between women using mostly cups or mostly tampons (Table 1).

**Table 1:** Association between menstrual cups usage in comparison with tampons. Results show the odds ratio (OR) of the factors selected in the best generalised linear model against menstrual cup as the response variable and using an Akaike Information Criterion corrected for small sample size (AICc). SE stands for standard error and CI for confidence interval.

| Model [1]: Menstrual cup | OR | OR SE | CI 2.5% | CI 95% | p.value |  |
| --- | --- | --- | --- | --- | --- | --- |
| Urinary tract infection | 3.45 | 2.37 | 0.86 | 13.37 | 0.07 | • |
| Smoking | 0.18 | 0.10 | 0.06 | 0.49 | 0.001 | ** |
| HPV positive (focal) | 0.36 | 0.18 | 0.13 | 0.92 | 0.04 | * |
| Number of partners over the last 12 months | 1.04 | 0.02 | 1.00 | 1.08 | 0.04 | * |
| Fungal infection | 6.55 | 4.22 | 1.86 | 24.10 | 0.004 | ** |

\*\* $p < 0.01$ ; \* $p < 0.05$ ; • $p < 0.1$

The demographic, behavioural, and biological analyses were performed on the first two visits of the participant (V1 and V2), which were spread 4 weeks apart.

### Patient involvement

Women participating in the study benefited from free gynaecological consults and from a compensation fee, as detailed in the study protocol [13]. Furthermore, two information leaflets on HPV infections in young women and on the vaginal microbiota were written in collaboration with the University Hospital of Montpellier and handed out to study participants upon inclusion. Finally, participants who have finished the follow-up and who accept to be informed by the scientific results of the study receive information by email.

### Biological analyses

Antibodies were analysed according to methods from Waterboer et al. [14] and already partly analysed in Murall et al. [15] in the context of HPV infection.

The cytokine data were obtained using MesoScale discovery (MSD) technology from vaginal secretions collected using ophthalmic sponges as described in Murall et al. [13] and was already analysed in the context of HPV infections by Selinger et al. [16]. We used the same protocol to obtain normalised values (per total protein concentration).

The microbiota profiling was performed via metabarcoding using the V3-V4 region of the 16S gene, as discussed in the study protocol [13]. The assignment of the community state type was done using the VALENCIA software package [17].

### Statistical analysis

We used binomial regression models for models in Table 1, Figure 1A, and in Extended data Tables S4 and S5. For each model, we computed the odds ratios associated with each predictor along with a 95% confidence interval. All analyses were performed in R (4.1.2) [18]. We used the glmulti function to conduct binomial regressions and selected the best model using the Akaike Information Criterion corrected for small sample sizes (AICc). We used a lowest AICc +5 interval for the most probable best models (Table S3 and S4) [19]. For clustering analysis, factor analysis of mixed data (FAMD) was used when combining both factor and numeric data (Figure 1F), and multiple correspondence analysis (MCA) was used when analysing binary data (Figure S2) [20].

**Figure 1:**
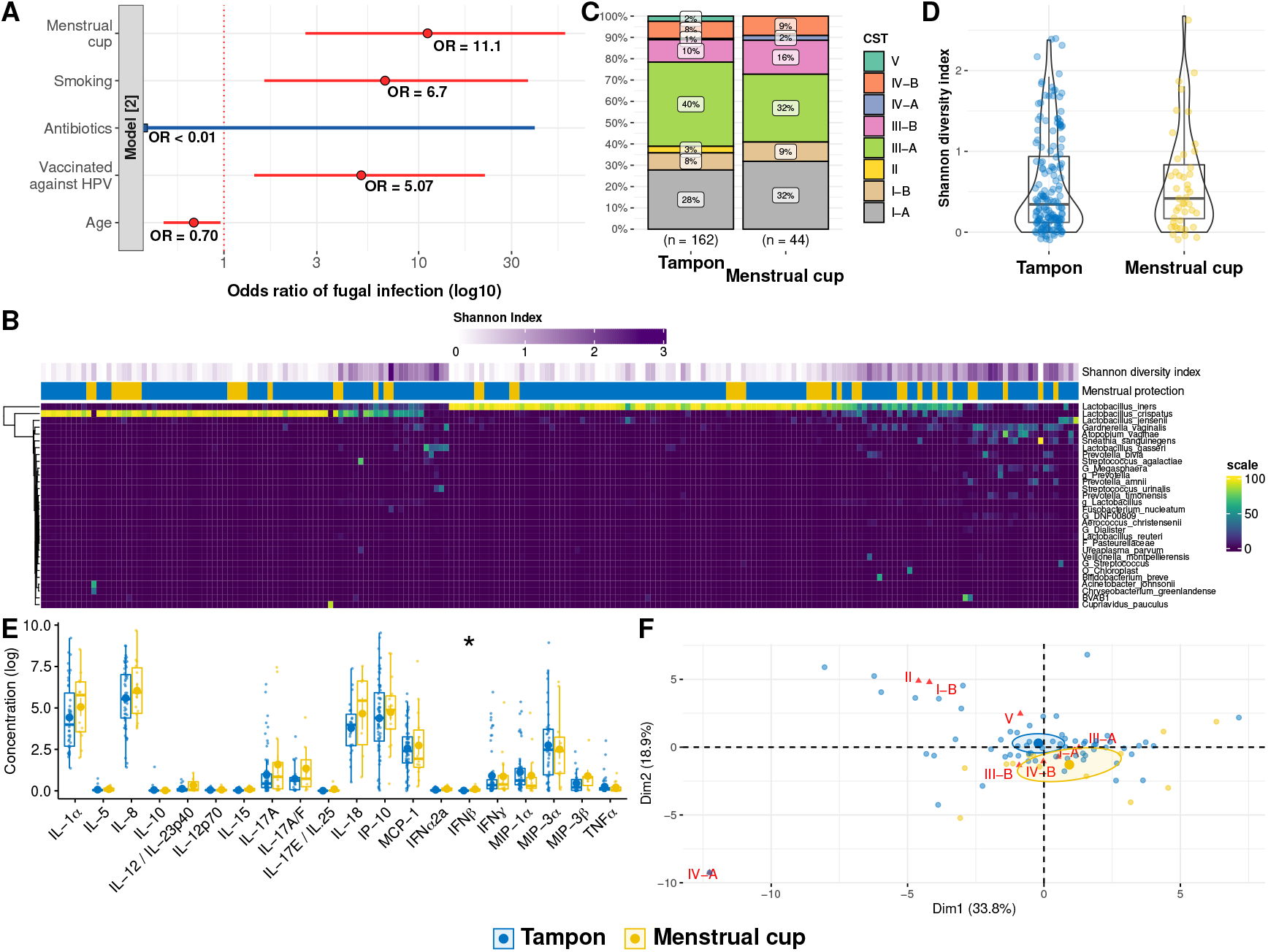
Epidemiological, microbiological, and immunological differences between women using mainly menstrual cups or tampons. A) Odds ratio of covariates associated with the risk of fungal infection detection, B) Abundance and diversity of the main bacterial species found in participants, C) Community State Types (CST) distribution, D) Shannon diversity index, E) Cytokines local concentrations (log), and F) Outcome of a multi-parametric clustering analysis using factor analysis of mixed data (FAMD). In A, red indicates significance (*p <* 0.05) and blue indicates non-significance. In D, E, and F, colours show the type of menstrual product used (tampons in blue and cups in yellow). In F, the CSTs are shown in red.

### Core outcome set

No core outcome set (COS) was used in the study. According to the CROWN database, the only potential relevant COS could have been “Heavy menstrual bleeding” but we found no general COS about menstrual health.

## Data availability

The data that support the findings of this study are available from the corresponding author upon request, and from the Zenodo public repository (XXXXX) upon publication.

## Results

The 103 women who could be included in the analysis were aged from 18 to 25 years old and primarily university students. Their main demographic characteristics are shown in Table S1. We stratified the population according to the most frequent type of menstrual product used, *i*.*e*. either tampons, *n* = 81 women, or menstrual cups, *n* = 22 (see the Supplementary Methods). The self-reported stress level was the only variable for which the two groups differed slightly (*p − value* = 0.08), with menstrual cup users reporting more frequently a moderate level of stress than tampon users (Supplementary Table S1).

We used generalised linear models to detect potential differences in key covariates (listed in Supplementary Table 2) between the two groups. The best model, according to our selection approach (see Supplementary Methods), shows that a significantly lower fraction of menstrual cup users identifies themselves as smokers compared to tampon users (*OR* = 0.18) and that the fraction of women who report using menstrual cups presenting a genital infection by human papillomaviruses (HPVs) is also lower (*OR* = 0.36). Conversely, reporting using menstrual cups was more associated with reporting a recent fungal genital infection (*OR* = 6.55). There was also a marginally significant trend of increased risk of urinary tract infection (Table 1). Importantly, Smoking and Fungal infection were the only two covariates to be present and significant in the best model and in the models comparable to it based on an Akaike Information Criterion (see Supplementary Methods and Table 2).

**Table 2:** Proportion of covariables presence among the 93 best models (Model 1) selected by AICc (*AICc* + 5)

| **Model [1]: Menstrual cup** | Presence in best<br>models (#) | Presence in best<br>models (%) | p.value<br>< 0.1(%) | p.value<br>< 0.05(%) |
| --- | --- | --- | --- | --- |
| Smoking | 93 | 100 | 100 | 100 |
| Fungal infection | 93 | 100 | 100 | 100 |
| HPV positive (focal) | 72 | 77.42 | 73.12 | 45.16 |
| Number of partners over the last 12 months | 71 | 76.34 | 46.24 | 21.51 |
| Urinary tract infection | 53 | 56.99 | 37.63 | 0 |
| Age | 40 | 43.01 | 19.35 | 0 |
| Menses | 19 | 20.43 | 0 | 0 |
| HPV positive (multiple) | 20 | 21.51 | 0 | 0 |
| Lubricant | 19 | 20.43 | 0 | 0 |
| Intercourse with an occasional partner (last week) | 14 | 15.05 | 0 | 0 |
| Intercourse with a regular partner (last week) | 13 | 13.98 | 0 | 0 |
| Vaccinated against HPV | 16 | 17.2 | 0 | 0 |
| Stress level (1) | 2 | 2.15 | 0 | 0 |
| Stress level (2) | 2 | 2.15 | 0 | 0 |
| Stress level (3 (Max)) | 2 | 2.15 | 0 | 0 |

To better understand the association between the type of menstrual products used and the occurrence of fungal infections, we performed another set of generalised linear models, this time evaluating the covariates associated with reporting a fungal infection. The selection procedure yielded a (best) model according to which smoking (p = 0.02) and the use of menstrual cups (*p* = 0.002) were associated with a higher reporting of recent fungal infection (Figure 1A). There was also a significant association between being vaccinated against HPV and reporting a recent fungal infection (*p* = 0.02). Of note, among women aged 18 to 25 years, reporting of fungal infections was less common in older cohorts. (*p* = 0.03). The type of menstrual product used the most (cups or tampons) was the only covariate present and significant in all of the 124 next best models selected (Supplementary Table S3), further confirming its association with reporting fungal infections in our cohort.

To study a potential association between the type of menstrual protection used and the vaginal microbiota, we performed a 16S metabarcoding analysis (Figure 1B). We found no significant difference in community state types (CST) composition (Figure 1C), although there were some qualitative differences. For instance, none of the menstrual cup users displayed a CST II or V [21]. We also did not find any significant differences in microbiota diversity, assessed using Shannon diversity index, between women using menstrual cups or tampons (Figure 1D).

To assess the potential effect of menstrual cups on the local immune response, we analysed cytokines and chemokine relative concentrations in cervical samples [16]. Among the 20 analytes measured, only IFN-β appeared to be significantly increased in women using cups (*p* = 0.044), although this association did not withstand correction for multiple testing comparisons (*p* = 0.89) (Figure 1E).

Finally, we performed a profile analysis using a factor analysis of mixed data (FAMD) approach with CSTs distribution, Shannon diversity index, cytokines, and chemokines relative concentrations. The results show that women who use tampons and women using menstrual cups fall into different clusters (Figure 1F and Supplementary Figure S1), suggesting that the type of menstrual product used could be shaping the local immunological and microbial environment. Conversely, a similar multiple correspondence analysis (MCA) approach using blood seropositivity status for IgG and IgM of several sexually transmitted infections (STIs), including HPVs (see Supplementary Methods) detected no clustering effect (Supplementary Figure S2), hinting that the women in these two groups originate from similar populations in terms of the risk of exposition to STIs.

## Discussion

Menstrual cups are gaining in popularity as an environmentally sustainable and affordable type of menstrual protection [10]. However, there currently is a lack of epidemiological and biological data to assess their safety. Using the PAPCLEAR study [13], we compared microbiological, immunological, and epidemiological profiles of women depending on whether they report using mainly cups or tampons.

### Main findings

We did not identify strong demographic or behavioural biases between our two populations, except for reported stress levels. On the epidemiological side, we detected a strong association between reporting using menstrual cups and recent fungal genital infections. We also found a marginally significant positive association with urinary tract infections. Finally, we identified a negative significant association with HPV infection. Note that women using menstrual cups reported a greater number of partners and less smoking than women using tampons, which suggests the decreased HPV prevalence is not due to differences in sexual contact. To further understand the occurrence of fungal infections, we performed another model which confirmed their strong association with menstrual cup use, as well as smoking, number of partners, and vaccination against HPV.

Biological analyses revealed an absence of significant difference between vaginal microbiota compositions depending on the type of menstrual product used. For local cytokine and chemokine relative concentrations, only one potential difference was identified in IFN-*β*. However, the joint analysis of microbiota and immunological data showed that women segregate into two clusters based on the type of menstrual product they use most. Similar analyses using circulating antibodies found no such clustering patterns, which reinforces our conclusion that the two populations of women studied do not differ from an epidemiological standpoint (their past exposition to STIs being comparable) but, potentially, from their local vaginal environment.

### Strengths and limitations

Our study has several limitations, the strongest being the relatively small sample size of the cohort used (*n* = 103), which was originally designed to study HPV infections. This may hinder our ability to detect moderate or subtle changes induced by the menstrual cups in the vaginal environment. Another limitation of the study lies in its cross-sectional nature. Further longitudinal analyses would be helpful to establish long-term potential impacts of the use of menstrual cups on the local environment, for example, on the vaginal microbiota composition. Finally, we cannot provide more details regarding the fungal species or their abundance. From a statistical standpoint, a potential issue has to do with pseudo-replication since we analyse two visits for each participant. To address it, we analysed each of the two visits independently. In both cases, the associations between menstrual cup use, smoking, and fungal infection were at least significant (Table S4 and S5). Interestingly, for the second visit, the time frame for reporting a fungal infection (since the last visit, i.e. one month) was smaller than for the inclusion visit (three months) and the effect is stronger in the former case.

Finally, other factors have been shown to shape the vaginal environment [21]. Here, we included for instance the use of lubricants in the analysis but more detailed studies could also include contraception methods or probiotic use.

### Interpretation

Earlier work on menstrual cup use and women’s health is limited and summarised in a recent meta-analysis [10]. Our results are consistent with earlier studies, which do not show adverse effects on the vaginal microbiota. Unfortunately, this earlier metaanalysis studied infections in general and it would require a new meta-analysis to focus on fungal infections in particular. However, the meta-analysis reported nine cases of urinary tract complaints and we do find that menstrual cups users reported more urinary tract infections than tampons users, although to a lesser extent than fungal infections.

## Conclusion

Menstrual cups are one of the most popular alternatives to disposable menstrual products. As the demand for ‘eco-friendly’ and less expensive menstrual products arises, it is crucial to better understand the effect of these products on women’s health. Our findings can help shape public health policies regarding the use of menstrual cups and underline the need for additional studies to be conducted that incorporate epidemiological, clinical, and biological outcomes.

## Data Availability

Table 1, 2 and Figure 1, as well as Extended Data Tables S1, S3, S4, S5 and Figure S2 have
associated raw data. The data that support the findings of this study are available from the corresponding author upon request, and data are available in the Zenodo public repository (XXXXX). 

## Acknowledgements

The authors thank all participants of the PAPCLEAR study and clinical staff and nurses for their help.

## Contributions

NT, CLM, NBo, and SA designed the study. NT, CLM, BE, BE, TK, IGB, and SA designed the experiments. CB, VB, SGro, MR, and NBe performed experiments. CH, JRa, and TW contributed reagents, materials, and analysis tools. CB, VB, SGra, SGro, MR, MB, CG, VT, VF, CLM, JRe, IGB, MSe, NBo, and SA contributed to study design, patient recruitment, and clinical data acquisition. NT, IBU, BE, BR, CS, TK, CLM, and SA performed data analyses. NT, IBU, IGB, CLM, and SA wrote the initial version of the manuscript. All authors approved the final version of the manuscript.

## Ethics

The PAPCLEAR trial is promoted by the Centre Hospitalier Universitaire (CHU) de Montpellier and has been approved by the Comité de Protection des Personnes (CPP) Sud Méditerranée I on 11 May 2016 (CPP number 16 42, reference number ID RCB 2016-A00712-49); by the Comité Consultatif sur le Traitement de l’Information en matière de Recherche dans le domaine de la Santé on 12 July 2016 (reference number 16.504); and by the Commission Nationale Informatique et Libertés on 16 December 2016 (reference number MMS/ABD/AR1612278, decision number DR-2016–488). This trial was authorised by the Agence Nationale de Sécurité du Médicament et des Produits de Santé on 20 July 2016 (reference 20160072000007). The ClinicalTrials.gov identifier is NCT02946346. All participants provided written informed consent.

## Bibliography

1. Hennegan J, Winkler IT, Bobel C, Keiser D, Hampton J, Larsson G, Chandra-Mouli V, Plesons M, and Mahon T. Menstrual health: a definition for policy, practice, and research. Sex. Reprod. Health Matters 2021; 29. doi: 10.1080/26410397.2021.1911618

2. Carneiro MM. Menstrual poverty: enough is enough. Women Health 2021; 61:721–2. doi: 10.1080/03630242.2021.1970502

3. Phillips-Howard PA, Nyothach E, Kuile FO ter, Omoto J, Wang D, Zeh C, Onyango C, Mason L, Alexander KT, Odhiambo FO, Eleveld A, Mohammed A, Eijk AM van, Edwards RT, Vulule J, Faragher B, and Laserson KF. Menstrual cups and sanitary pads to reduce school attrition, and sexually transmitted and reproductive tract infections: a cluster randomised controlled feasibility study in rural Western Kenya. BMJ Open 2016; 6. doi: 10.1136/bmjopen-2016-013229

4. Torondel B, Sinha S, Mohanty JR, Swain T, Sahoo P, Panda B, Nayak A, Bara M, Bilung B, Cumming O, Panigrahi P, and Das P. Association between unhygienic menstrual management practices and prevalence of lower reproductive tract infections: a hospital-based cross-sectional study in Odisha, India. BMC Infect Dis 2018; 18:473

5. Smith SB and Ravel J. The vaginal microbiota, host defence and reproductive physiology. J Physiol 2017; 595:451–63. doi: 10.1113/JP271694

6. McKinnon LR, Achilles SL, Bradshaw CS, Burgener A, Crucitti T, Fredricks DN, Jaspan HB, Kaul R, Kaushic C, Klatt N, Kwon DS, Marrazzo JM, Masson L, McClelland RS, Ravel J, Wijgert Jhhm van de, Vodstrcil LA, and Tachedjian G. The Evolving Facets of Bacterial Vaginosis: Implications for HIV Transmission. AIDS Res Hum Retroviruses 2019; 35:219–28. doi: 10.1089/aid.2018.0304

7. Ma B, France MT, Crabtree J, Holm JB, Humphrys MS, Brotman RM, and Ravel J. A comprehensive non-redundant gene catalog reveals extensive within-community intraspecies diversity in the human vagina. Nat Commun 2020; 11:940

8. Zhou JZ, Way SS, and Chen K. Immunology of the Uterine and Vaginal Mucosae. Trends Immunol 2018 Apr; 39:302–14

9. Wira CR, Rodriguez-Garcia M, and Patel MV. The role of sex hormones in immune protection of the female reproductive tract. Nat Rev Immunol 2015 Apr; 15:217– 30

10. Eijk AM van, Zulaika G, Lenchner M, Mason L, Sivakami M, Nyothach E, Unger H, Laserson K, and Phillips-Howard PA. Menstrual cup use, leakage, acceptability, safety, and availability: a systematic review and meta-analysis. Lancet Public Health 2019; 4:e376–e393. doi: 10.1016/S2468-2667(19)30111-2

11. North BB and Oldham MJ. Preclinical, clinical, and over-the-counter postmarketing experience with a new vaginal cup: menstrual collection. J Womens Health 2011; 20:303–11. doi: 10.1089/jwh.2009.1929

12. Nonfoux L, Chiaruzzi M, Badiou C, Baude J, Tristan A, Thioulouse J, Muller D, Prigent-Combaret C, and Lina G. Impact of Currently Marketed Tampons and Menstrual Cups on Staphylococcus aureus Growth and Toxic Shock Syndrome Toxin 1 Production In Vitro. Appl. Environ. Microbiol. 2018; 84. doi: 10.1128/AEM.00351-18

13. Murall CL, Rahmoun M, Selinger C, Baldellou M, Bernat C, Bonneau M, Boué V, Buisson M, Christophe G, D’Auria G, Taroni FD, Foulongne V, Froissart R, Graf C, Grasset S, Groc S, Hirtz C, Jaussent A, Lajoie J, Lorcy F, Picot E, Picot MC, Ravel J, Reynes J, Rousset T, Seddiki A, Teirlinck M, Tribout V, Tuaillon É, Waterboer T, Jacobs N, Bravo IG, Segondy M, Boulle N, and Alizon S. Natural history, dynamics, and ecology of human papillomaviruses in genital infections of young women: Protocol of the PAPCLEAR cohort study. BMJ Open 2019; 9:25129. doi: 10.1136/bmjopen-2018-025129

14. Waterboer T, Sehr P, Michael KM, Franceschi S, Nieland JD, Joos TO, Templin MF, and Pawlita M. Multiplex human papillomavirus serology based on in situ-purified glutathione s-transferase fusion proteins. Clin Chem 2005; 51:1845–53

15. Murall CL, Reyné B, Selinger C, Bernat C, Boué V, Grasset S, Groc S, Rahmoun M, Bender N, Bonneau M, Foulongne V, Graf C, Picot E, Picot MC, Tribout V, Waterboer T, Bravo IG, Reynes J, Segondy M, Boulle N, and Alizon S. HPV cervical infections and serological status in vaccinated and unvaccinated women. Vaccine 2020; 38:8167–74. doi: 10.1016/j.vaccine.2020.10.078

16. Selinger C, Rahmoun M, Murall CL, Bernat C, Boué V, Bonneau M, Graf C, Grasset S, Groc S, Reynes J, Hirtz C, Jacobs N, and Alizon S. Cytokine response following perturbation of the cervicovaginal milieu during HPV genital infection. Immunol. Res. 2021; 69:255–63. doi: 10.1007/S12026-021-09196-2

17. France MT, Ma B, Gajer P, Brown S, Humphrys MS, Holm JB, Waetjen LE, Brotman RM, and Ravel J. VALENCIA: a nearest centroid classification method for vaginal microbial communities based on composition. Microbiome 2020; 8:166. doi: 10.1186/s40168-020-00934-6

18. R Core Team. R: A Language and Environment for Statistical Computing. R Foundation for Statistical Computing. Vienna, Austria, 2021. Available from: https://www.R-project.org/

19. Burnham K and Anderson D. Model Selection and Inference: A Practical Information-Theoretic Approach. 2. 2002 :75

20. Lê S, Josse J, and Husson F. FactoMineR: An R Package for Multivariate Analysis. Journal of Statistical Software 2008; 25:1–18. doi: 10.18637/jss.v025.i01

21. Ravel J, Gajer P, Abdo Z, Schneider GM, Koenig SS, McCulle SL, Karlebach S, Gorle R, Russell J, Tacket CO, Brotman RM, Davis CC, Ault K, Peralta L, and Forney LJ. Vaginal microbiome of reproductive-age women. Proc Nat Acad Sci USA 2011; 108:4680–7. doi: 10.1073/pnas.1002611107

