## Supporting information for "Increased risk of fungal infection detection in women using menstrual cups vs. tampons: a cross-sectional study"

January 6, 2022

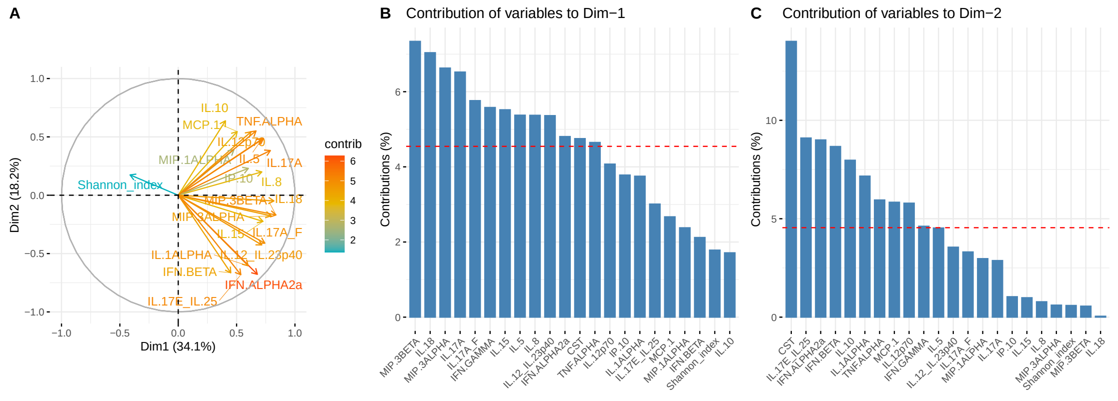


Figure S1: **Contributions of covariates to the FAMD analysis in Figure 1**
**A:** Graph of variables vectors and their relative contribution to the FAMD analysis in Figure 1F. **B and C:** Percentage of contributions to dimension 1 (**A**) and dimension 2 (**B**) of the FAMD analysis in Figure 1F)

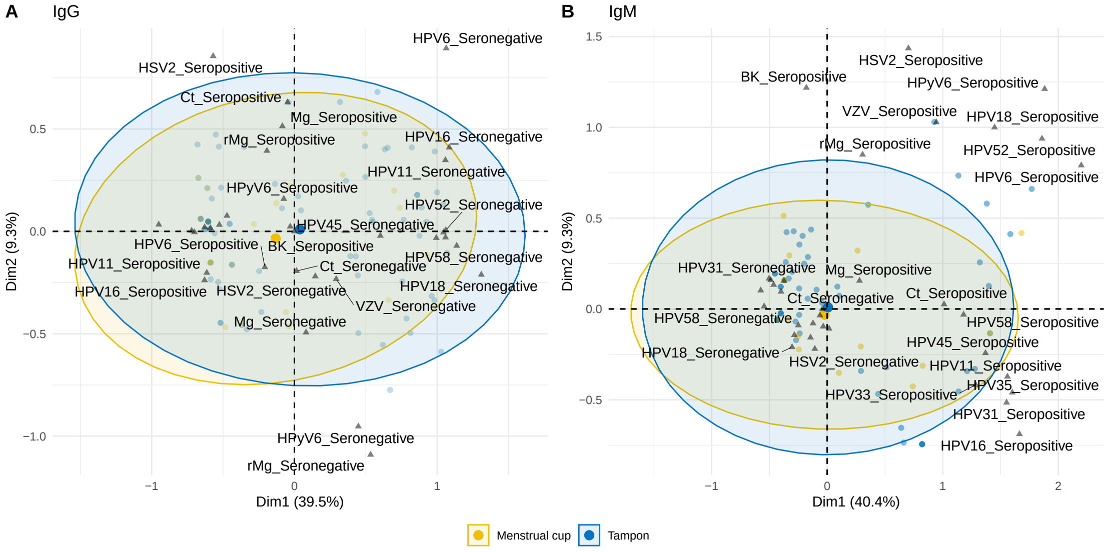


Figure S2: **Multiple Correspondance Analysis of seropositivity for diverse STI antibodies**

**A** IgG and **B** IgM seropositivity for a panel of STI antibodies. Antibodies included are the following. Human papillomarivus: (anti-HPV6, anti-HPV11, anti-HPV16, anti-HPV18, anti-HPV31, anti-HPV33, anti-HPV35, anti-HPV45, anti-HPV52 and anti-HPV58); Human polyomavirus: (anti-BK and anti-HPyV6), Human Herpes simplex virus: (anti-HSV2 and anti-VZV); *Chlamydia trachomatis:* (anti-Ct) and *Mycoplasma genitalium:* (anti-Mg and anti-rMg).

Table S1: Key characteristics of participants included in the study. $n$ indicates the number of individuals, p-value refers to the outcome of a Kruskal-Wallis Rank Sum Test (`kruskal.test` function in R), IQR is the interquartile range, SD the standard deviation.

| | Tampon | Menstrual cup | p-value |
|---|---|---|---|
| $n$ | 81 | 22 | |
| Follow-up duration (days, median [IQR]) | 301.00 [203.00, 455.00] | 409.50 [238.50, 515.50] | 0.165 |
| Number of partners over the last 12 months (median [IQR]) | 8.00 [3.00, 15.00] | 6.00 [4.00, 14.75] | 0.886 |
| Age (years, median [IQR]) | 21.30 (1.85) | 21.95 (2.50) | 0.174 |
| Age at menarchy (years, median [IQR]) | 13.00 [12.00, 14.00] | 13.00 [12.25, 14.00] | 0.665 |
| Body Mass Index (BMI, median [IQR]) | 21.03 [19.61, 23.34] | 21.77 [20.66, 23.41] | 0.387 |
| Antibiotics = Yes (%) | 5 ( 6.2) | 2 ( 9.1) | 0.996 |
| Smoking = Yes (%) | 31 (40.3) | 5 (22.7) | 0.209 |
| Lubricant (%) | 28 (35.0) | 8 (36.4) | 1.000 |
| HPV positive (positive for the same HPV at the two visits) (%) | 40 (49.4) | 7 (31.8) | 0.220 |
| Intercourse with a regular partner (last week) | 50 (61.7) | 13 (59.1) | 1.000 |
| Intercourse with an occasional partner (last week) | 11 (13.6) | 3 (13.6) | 1.000 |
| Stress level (%) | | | 0.081 |
| 0 (Min) | 17 (21.0) | 4 (18.2) | |
| 1 | 35 (43.2) | 7 (31.8) | |
| 2 | 20 (24.7) | 11 (50.0) | |
| 3 (Max) | 9 (11.1) | 0 ( 0.0) | |
| Self-reported menses during the past week | 44 (54.3) | 15 (68.2) | 0.356 |
| Co-infected by multiple HPVs | 27 (33.3) | 7 (31.8) | 1.000 |
| Age at sexual debut (mean (SD)) | 16.40 (1.55) | 16.50 (1.97) | 0.792 |
| HPV vaccinated (%) | 37 (47.4) | 14 (70.0) | 0.121 |

Table S2: Factors used in the best models selection by AICc.

| Covariate | Data type | Values | Description | source |
|---|---|---|---|---|
| Number of partners over the last 12 months | Numeric | 1 to 63 | Number of partners over the last 12 months | Questionnaire at inclusion visit |
| Smoking | Binary | Yes/No | Self-reported smoking during past week | Questionnaire at each visit |
| Intercourse with an occasional partner | Binary | Yes/No | Intercourse with an occasional partner during the past week | Questionnaire at each visit |
| HPV vaccinated | Binary | Yes/No | Vaccinated against HPV | Questionnaire at inclusion visit |
| Menstrual protection | Binary | Menstrual cups/ Tampons | Questionnaire at each visit | |
| HPV focal | Binary | Yes/No | DEIA positive for at least one HPV genotype at two consecutive visits | Laboratory testing |
| Age | Numeric | 18:25 | Self-reported current age | Questionnaire at inclusion visit |
| Lubricant | Binary | Yes/No | Self-reported using sexual lubricant during past week | Questionnaire at each visit |
| HPV positive (multiple HPV) | Binary | Yes/No | DEIA positive for at more than one HPV genotype | Laboratory testing |
| Antibiotics | Binary | Yes/No | Used antibiotics during the past week | Questionnaire at each visit |
| Intercourse with a regular partner | Binary | Yes/No | Self-reported intercourse with a regular partner during past week | Questionnaire at each visit |
| Menses | Binary | Yes/No | Self-reported menses during past week | Questionnaire at each visit |
| Stress level | Categorical | 0 (Min), 1, 2, 3 (Max) | Self-reported stress level during past week | Questionnaire at each visit |
| Recent urinary tract infection | Binary | Yes/No | Self-reported (over the last 3 months at inclusion and since the last visit later) | Questionnaire at each visit |
| Recent fungal infection | Binary | Yes/No | Self-reported (over the last 3 months at inclusion and since the last visit later) | Questionnaire at each visit |

| **Model [2]: Fungal infection** | Presence in best models (#) | Presence in best models (%) | p.value < 0.1(%) | p.value < 0.05(%) |
|---|---|---|---|---|
| Menstrual cup | 124 | 100 | 100 | 100 |
| Smoking | 124 | 100 | 100 | 96.77 |
| Vaccinated against HPV | 107 | 86.29 | 86.29 | 66.13 |
| Age | 61 | 49.19 | 22.58 | 16.94 |
| Number of partners over the last 12 months | 87 | 70.16 | 33.87 | 1.61 |
| Antibiotics | 118 | 95.16 | 0 | 0 |
| Intercourse with a regular partner (last week) | 38 | 30.65 | 0 | 0 |
| Lubricant | 40 | 32.26 | 0 | 0 |
| Intercourse with an occasional partner (last week) | 34 | 27.42 | 0 | 0 |
| HPV positive (focal) | 28 | 22.58 | 0 | 0 |
| HPV positive (multiple) | 32 | 25.81 | 0 | 0 |
| Menses | 24 | 19.35 | 0 | 0 |
| Stress level (1) | 11 | 8.87 | 0 | 0 |
| Stress level (2) | 11 | 8.87 | 0 | 0 |
| Stress level (3 (Max)) | 11 | 8.87 | 0 | 0 |

Table S3: Proportion of covariables presence among the 124 best models (Model 1) selected by AICc (AICc + 5)

Table S4: Best model selected for covariates associated with menstrual cups usage in comparison with tampons - **First visit**

| **Model [1]: Menstrual cup** | OR | OR SE | CI 2.5% | CI 95% | p.value | |
|---|---|---|---|---|---|---|
| Smoking | 0.06 | 0.06 | 0.01 | 0.34 | 0.00 | ** |
| Stress level (1) | 0.38 | 0.43 | 0.04 | 4.01 | 0.40 | |
| Stress level (2) | 4.86 | 5.51 | 0.63 | 62.01 | 0.16 | |
| Stress level (3 (Max)) | 0.00 | 0.00 | 0.00 | NA | 0.99 | |
| Fungal infection | 8.47 | 10.25 | 0.83 | 114.89 | 0.08 | • |

$^*p < 0.05$; • $p < 0.1$

Table S5: Best model selected for covariates associated with menstrual cups usage in comparison with tampons - **Second visit**

| **Model [1]: Menstrual cup** | OR | OR SE | CI 2.5% | CI 95% | p.value | |
|---|---|---|---|---|---|---|
| Smoking | 0.12 | 0.11 | 0.01 | 0.61 | 0.02 | * |
| HPV positive (focal) | 0.31 | 0.22 | 0.07 | 1.15 | 0.09 | • |
| Age | 1.33 | 0.22 | 0.97 | 1.88 | 0.09 | • |
| Number of partners over the last 12 months | 1.04 | 0.03 | 0.99 | 1.11 | 0.12 | |
| Fungal infection | 9.73 | 8.96 | 1.66 | 68.30 | 0.01 | * |

$^*p < 0.05$; • $p < 0.1$
